# Circulating Extracellular Vesicle miRNAs for Distinguishing Prostate Cancer and Benign Prostatic Hyperplasia

**DOI:** 10.64898/2026.02.18.26346569

**Authors:** Ting Ding, Xin Zhang, Lijuan Yu

**Affiliations:** Department of Laboratory Medicine, The Second Hospital Affiliated to Xi’an Jiaotong University, Xi’an Jiaotong University, Xi’an, 710049, China; Department of Laboratory Medicine, Xijing Hospital, Fourth Military Medical University (Air Force Medical University), Xi’an 710032, China; Department of Laboratory Medicine, Guangdong Provincial Key Laboratory of Precision Medical Diagnostics, Guangdong Engineering and Technology Research Center for Rapid Diagnostic Biosensors, Guangdong Provincial Key Laboratory of Single-cell and Extracellular Vesicles, Nanfang Hospital, Southern Medical University, Guangzhou, 510515, P. R. China; Sahlgrenska Center for Cancer Research, Department of Surgery, Institute of Clinical Sciences, Sahlgrenska Academy, University of Gothenburg, Gothenburg, Sweden

**Keywords:** Prostate cancer, extracellular vesicles, miRNA, diagnosis

## Abstract

Our previous studies identified three microRNAs (miR-92a-1-5p, miR-375 and miR-148a-3p) potentially associated with prostate cancer (PCa), particularly in advanced stages such as bone-metastatic PCa. To evaluate their clinical diagnostic utility, we isolated extracellular vesicles (EVs) from the plasma of patients with benign prostatic hyperplasia (BPH) and PCa (including localized and bone-metastatic disease). The absolute quantification of these three miRNAs within plasma EVs was performed using digital PCR. Results indicated that miR-148a-3p alone possessed a good ability to discriminate between PCa and BPH. Notably, a combined panel of all three miRNAs demonstrated improved diagnostic performance, achieving an area under the curve (AUC) of 0.736 for distinguishing PCa from BPH. These findings suggest that the plasma EV-derived miRNA panel (miR-92a-3p, miR-148a-3p, and miR-375-3p) holds promise as an auxiliary diagnostic biomarker for PCa and may aid in identifying bone metastasis.

## INTRODUCTION

Prostate cancer (PCa) is a leading malignancy in men ^1^. A major and immediate clinical burden is the accurate differentiation of PCa from the highly prevalent benign prostatic hyperplasia (BPH), as both conditions present similarly and commonly elevate serum prostate-specific antigen (PSA). PSA, the standard biomarker, lacks the specificity required for this critical distinction, frequently leading to unnecessary biopsies and diagnostic delays ^2^. This highlights an urgent need for novel, non-invasive biomarkers. Liquid biopsy, particularly the analysis of molecular cargo in circulating extracellular vesicles (EVs), represents a promising avenue to develop diagnostic tools capable of reliably distinguishing PCa from BPH ^3^, with the added potential to identify aggressive disease such as bone metastasis.

In the search for such novel biomarkers, EVs have garnered significant attention as a superior liquid biopsy substrate. EVs are membrane-bound nanoparticles stably present in body fluids like plasma, and their molecular cargo reflects the pathophysiological state of their parent cells^4^. Among these cargoes, microRNAs (miRNAs) are particularly promising. These small non-coding RNAs are key regulators of gene expression and are deeply involved in cancer processes, including the development and metastasis of PCa^3^. Critically, EV-encapsulated miRNAs are protected from degradation, and their expression profiles in plasma have been shown to effectively discriminate cancer patients from healthy individuals, as well as metastatic from non-metastatic disease in various cancers ^5,6^. These properties position EV-derived miRNAs as ideal candidates for developing the specific biomarkers needed to address the pressing clinical challenge of distinguishing PCa from BPH.

In our previous preliminary study, three miRNAs (miR-92a-1-5p, miR-375 and miR-148a-3p) were identified via high-throughput sequencing and bioinformatics analysis as being potentially associated with PCa progression, particularly in bone-metastatic disease. However, the translational clinical value of this three-miRNA panel, especially as a diagnostic biomarker derived from plasma EVs, remained unclear. It was unknown whether their detection could reliably differentiate PCa from BPH or indicate the presence of bone metastasis, nor was it known if a combined panel would outperform individual markers. Therefore, to bridge this gap, this study aimed to perform absolute quantification of these three miRNAs in plasma EVs from cohorts of patients with BPH, localized PCa, and bone-metastatic PCa using digital PCR. We sought to evaluate their individual and combined diagnostic efficacy for both distinguishing PCa from BPH and for identifying bone metastasis, with the goal of validating a novel non-invasive biomarker panel for potential clinical application.

## MATERIAL AND METHODS

### Patients

With informed consent from the patients and approval from the hospital’s ethics committee (NO. KY20212016-C-1), we recruited 66 patients at the Department of Urology, the First Affiliated Hospital of the Air Force Medical University. These patients were histologically confirmed to have no other concurrent malignancies and had not yet received any treatment. After enrollment, plasma samples were collected from the patients. The staging and risk stratification of patients with PCa in this study followed the classification outlined in the “ESMO Clinical Practice Guidelines for Prostate Cancer: Diagnosis, Treatment, and Follow-Up” and EAU Guidelines for prostate cancer ^7,8^.

### Plasma isolation

First, EDTA anticoagulated blood samples collected under fasting conditions were centrifuged at 3000 g for 10 minutes at room temperature within 4 hours to separate the plasma. Subsequently, the supernatant was collected after centrifugation at 10,000 g for 10 minutes at room temperature, aliquoted, and stored at -80°C.

### EVs isolation and characterization

Prostate plasma EVs were isolated from serum samples using the Exosome Isolation Kit (Yugongbiomed, Shanghai, China) according to the manufacturer’s protocols. Briefly, plasma was pre-centrifuged to remove cells and debris, then mixed with the isolation reagent and incubated at 4°C. After recovery of the EV pellet through centrifugation, the precipitates were further purified using the provided purification columns to ensure high purity of the isolated vesicles.

### EVs characterization

The morphology of EVs was analyzed using transmission electron microscopy (TEM) (Tecnai, USA), as previously described ^9^. To examine the size distribution and particle concentration of EVs, fractionated samples were diluted in PBS for nanoparticle tracking analysis (NTA) using a ZetaView instrument (Particle Metrix, Germany). Western blotting (WB), following previous literature^10^, was used to detect two EV-positive markers and one EV-negative marker: TSG101 (Abcam ab125011, 1:1000) and CD9 (Abcam ab263019, 1:1000). Calnexin was purchased from Proteintech (10427-2, 1:500 dilution).

### miRNA Detection

RNA was extracted from the isolated EVs using the miRNeasy Mini Kit (Qiagen, Hilden, Germany) following the manufacturer’s instructions, and then using the PrimeScript™ RT reagent Kit (Takara Bio, Shiga, Japan) according to the manufacturer’s instructions for cDNA synthesis. The absolute quantification of miRNA was subsequently performed using the naica® Crystal Digital PCR system (Stilla Technologies, Villejuif, France). The 25 μL reaction mixture consisted of 1× naica® EvaGreen PCR MIX (Stilla Technologies), 200 nM of each forward and reverse primer (RiboBio, Guangzhou, China), and 2 μL of the cDNA template. The mixture was loaded into the Sapphire Chips, and the chips were placed into the Geode device. The integrated program performed droplet generation followed by thermal cycling: initial denaturation at 95°C for 2 min, followed by 45 cycles of 95°C for 5 s and 60°C for 30 s ^11^. After amplification, the chips were transferred to the Prism reader for six-channel fluorescence detection. The raw data were analyzed using Crystal Miner software (Stilla Technologies) to calculate the absolute concentration of target miRNA, expressed as copies/μL.

### Statistical analysis

Statistical analyses were performed using GraphPad Prism (version 9.0). All data are presented as means ± standard deviation. Comparisons were performed using *t* test and one-way ANOVA as appropriate, *P* < 0.05 was considered statistically significant (*, *P* < 0.05; **, *P* < 0.01; ***, *P* < 0.001).

## RESULTS

### Isolation and characterization of plasma EVs

Plasma EVs were successfully isolated from all patient cohorts. Transmission electron microscopy revealed that the isolated particles exhibited the typical cup-shaped morphology characteristic of EVs (Figure 1a). Nanoparticle tracking analysis confirmed their size distribution, with a predominant peak around 120 nm (Figure 1b). Western blot analysis further validated the EV identity, demonstrating positive expression of the canonical EV markers CD9 and TSG101, while the endoplasmic reticulum contaminant marker Calnexin was absent (Figure 1c). Collectively, these data confirm the successful isolation of high-purity plasma EVs suitable for downstream miRNA analysis.

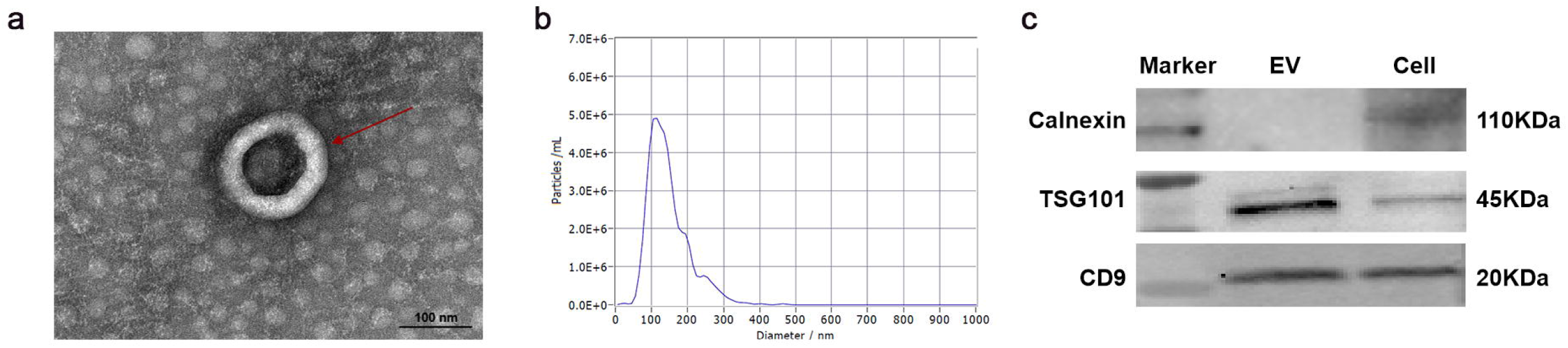

### Individual and Combined Diagnostic Performance of the miRNA Panel

To evaluate the diagnostic potential of the three miRNAs for prostate cancer, we first assessed their individual performance in distinguishing patients with PCa (including both localized and bone-metastatic disease) from those with BPH. While all three miRNAs displayed a trend of upregulation in the PCa group, only miR-148a-3p showed a statistically significant difference in expression between PCa and BPH (Figure 2a-c). Correspondingly, when analyzed individually via receiver operating characteristic (ROC) curves, none of the miRNAs achieved a satisfactory area under the curve (AUC) value for diagnosis (Figure 2d-f). Notably, the combined analysis of all three miRNAs (miR-92a-1-5p, miR-375, and miR-148a-3p) markedly improved diagnostic performance, yielding an AUC of 0.736 (Figure 2g) This result indicates that the synergistic use of this miRNA panel holds superior potential for distinguishing PCa from BPH compared to any single miRNA alone.

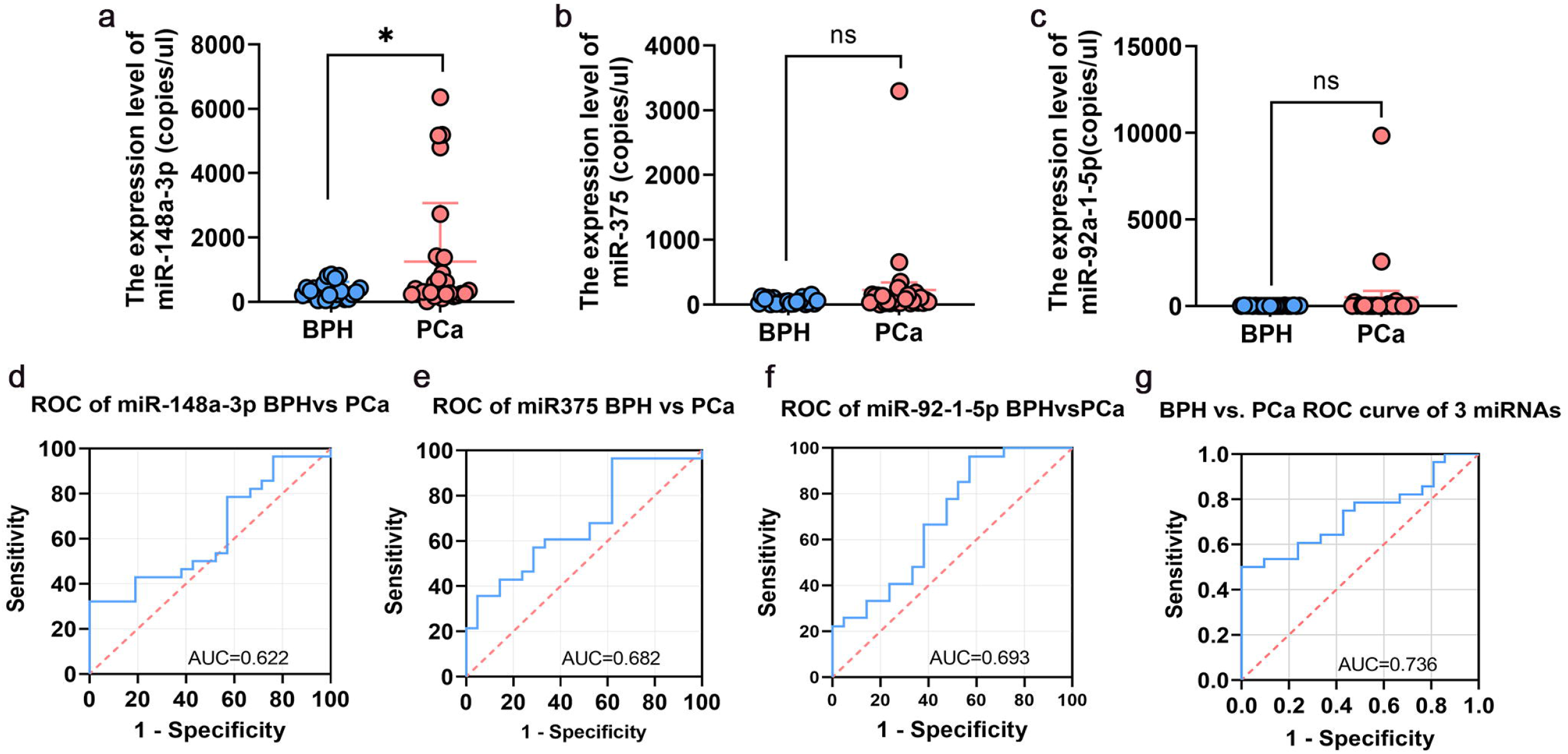

### Potential in differentiating bone metastatic PCa

We further evaluated the ability of the three miRNAs to identify bone-metastatic PCa by stratifying the cohort into three diagnostic groups: BPH, localized PCa, and bone-metastatic PCa. Analysis of individual miRNAs revealed that while all three were upregulated in localized PCa compared to BPH, their expression levels showed a slight downward trend in bone-metastatic cases (Supplementary Fig. 1a-c). Among them, miR-375 exhibited the most promising performance for distinguishing bone-metastatic PCa from BPH, yielding an AUC of 0.735 in ROC analysis (Supplementary Fig. 1d).

The combined diagnostic model based on the three-miRNA panel demonstrated enhanced discriminatory power. It achieved an AUC of 0.737 for distinguishing localized PCa from BPH (Fig. 3a) and an AUC of 0.766 for distinguishing bone-metastatic PCa from BPH (Fig. 3b). However, the model did not show significant diagnostic value for differentiating between localized PCa and bone-metastatic PCa (Fig. 3c), indicating its primary utility lies in distinguishing malignant from benign disease rather than in staging advanced PCa.

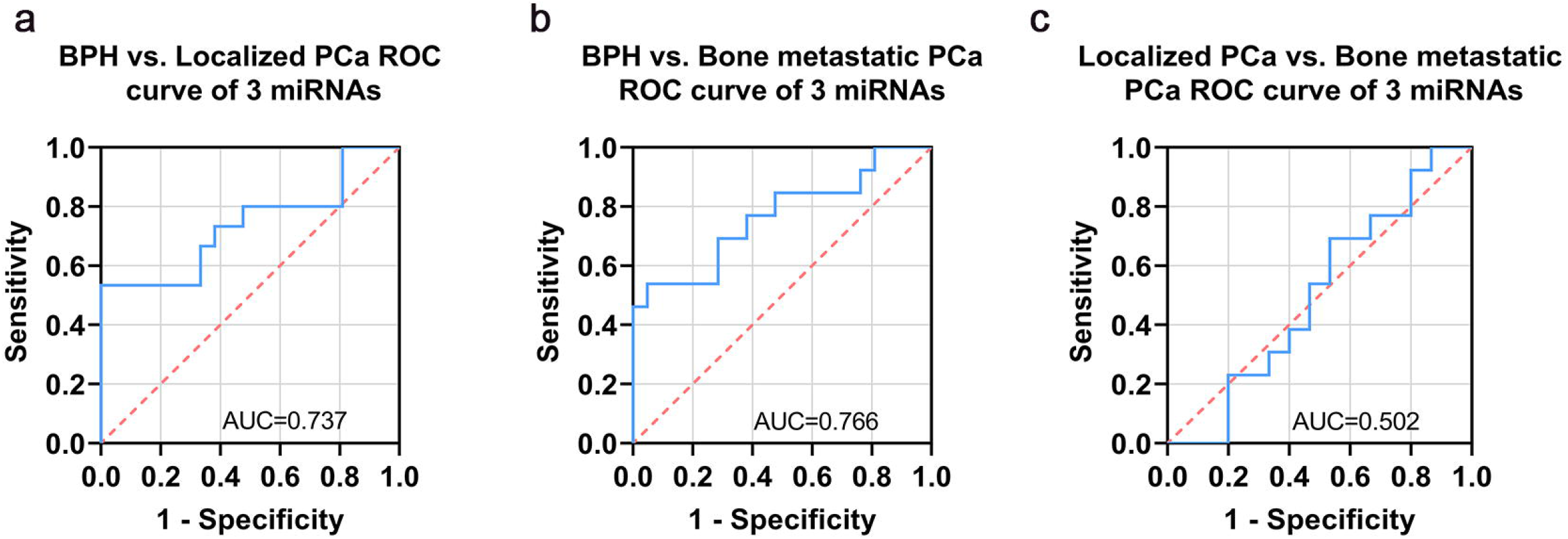

## DISCUSSION

While previous mechanistic studies, including our own, have implicated miRNAs such as miR□148a□3p, miR□92a□1□5p, and miR□375 in the pathogenesis of prostate cancer (PCa) ^12-14^, their translational value as clinically applicable biomarkers remain unclear. To bridge this gap, this study aimed to validate the diagnostic potential of a plasma EVs-derived miRNA panel, focusing on its ability to distinguish PCa from benign prostatic hyperplasia (BPH) and to identify bone-metastatic disease.

This study’s methodology is based on effectively isolating plasma EVs using a commercial kit. We confirmed the EVs’ typical morphology, size (∼120 nm), and marker expression (CD9, TSG101) without Calnexin contamination through electron microscopy, nanoparticle tracking, and western blot. This high-purity isolation is crucial for accurate miRNA analysis, ensuring reliable digital PCR results and strengthening our diagnostic conclusions.

Individually, miR-148a-3p, miR-92a-1-5p, and miR-375 lack sufficient diagnostic power to differentiate PCa from BPH. However, when combined, they achieve a significantly better AUC of 0.736, indicating the advantage of multi-miRNA panels over single markers. This combination may serve as a useful diagnostic tool alongside serum PSA testing, especially when PSA results are unclear, potentially reducing unnecessary biopsies. The study highlights the difficulty of identifying a single effective biomarker for PCa and supports the use of multiple biomarkers for more accurate diagnosis.

There are also some studies that have explored the value of these miRNAs in PCa. Several studies have confirmed that miR-92a is upregulated in PCa ^15-18^. However, these studies were all conducted on tissue or cell models. Our research is one of the first studies to demonstrate the diagnostic value of plasma exosomal miR-92a-1-5p for PCa. Regarding miR□148, Coradduzza et al. demonstrated its downregulation in PCa tissues and its role in cancer progression ^19,20^. Unfortunately, these studies only explored the value of miR-148 in the diagnosis of prostate cancer, but did not analyze its potential in differentiating PCa bone metastasis, and they detected miR-148a-3p in plasma rather than in EVs. For miR-375, numerous studies have confirmed its diagnostic value in PCa ^16,21-23^, and it has been found to be associated with the poor prognosis of PCa ^23-26^. However, most of these studies were conducted using tissue or cell models, and no study has explored the value of plasma EV miR-375 in PCa bone metastasis. Our study also found that miR□375 exhibits good diagnostic performance (AUC□=□0.735) for PCa bone metastasis detection from BPH, indicating its potential as a key biomarker. Literature supports that miR□375 promotes PCa cell migration and induces docetaxel resistance by targeting YAP1^24,26^. Moreover, exosomal miR□375 is significantly elevated in bone□metastatic tumors and contributes to the formation of a pre□metastatic niche in bone ^25^. These mechanisms likely underlie its high diagnostic value, linking its expression to bone□metastasis□related pathways.

Furthermore, our three-miRNA panel showed a complex pattern in staging: all miRNAs were higher in localized PCa than in BPH but decreased in bone-metastatic disease. This suggests a shift in EV-miRNA cargo during disease progression, possibly due to changes in tumor cell behavior or interactions with the bone microenvironment. The downregulation of these miRNAs in bone metastases may indicate a reprogramming of EV cargo by PCa cells, shifting from a “pro-growth” to a “niche adaptation” or “systemic crosstalk” role^12^. This dynamic change in miRNA profiles may explain why our panel, designed for malignancy detection, lacks specificity in distinguishing localized from metastatic disease, aligning with new insights into selective EV cargo sorting.

However, the model did not demonstrate effective diagnostic performance in distinguishing between localized PCa and bone-metastatic PCa. This limitation may stem from several factors. First, the relatively limited sample size in this study may affect the stability and statistical power of the model in subgroup analyses. Second, although the three selected miRNAs showed overall upregulation in malignant disease, their dynamic changes during disease progression may not sensitively reflect the molecular differences between localized progression and distant metastasis. Furthermore, the formation of bone metastasis in prostate cancer involves complex remodeling of the tumor microenvironment and systemic regulation; a single plasma EV miRNA profile may not fully capture the key biological characteristics of this process. Therefore, the current model is more suitable for assisting in the differentiation of benign and malignant prostate conditions, while it still has clear shortcomings in precise staging.

Future research could further optimize model construction by expanding sample sizes, incorporating novel biomarkers with greater staging specificity, and adopting multi-omics analysis strategies, thereby advancing its application in the precise diagnosis and stratified management of PCa.

Collectively, this study confirms that a three-miRNA signature (miR-148a-3p, miR-92a-1-5p, miR-375) from plasma extracellular vesicles can effectively differentiate prostate cancer from benign prostatic hyperplasia, with a combined AUC of 0.736. The panel also improves detection of bone-metastatic prostate cancer (AUC = 0.766) but lacks specificity for distinguishing localized from metastatic disease. These results underscore the potential of EV-based liquid biopsy and the need for further biomarker refinement for precise prostate cancer stratification.

## Supporting information

Graphic Abstract

## Data Availability

All data produced in the present study are available upon reasonable request to the corresponding author.

## DECLARATIONS

### Ethics approval

The research was conducted in accordance with the Declaration of Helsinki and was approved by the Animal Ethics Committee at Air Force Medical University, China.

### Consent for publication

The authors read and agree this version be published.

### Availability of data and materials

All the original data are available upon reasonable request for correspondence author.

## Acknowledgements

This work was supported by the Shaanxi Provincial Natural Science Basic Research Program to T.D. [under Grant No. 2025JC-YBQN-1185], the National Natural Science Foundation of China to L.Y. [under Grant No. 82203711] and the China Postdoctoral Science Foundation to L.Y. [under Grant No. 2021M701631].

## Conflict of interest statement

The authors declare no conflict of interest.

## Authors’ contributions

T.D. designed the experiments, performed the experiments, analyzed the data, wrote the draft manuscript and provided funds; X.Z revised the manuscript; L.Y. designed the experiments, coordinated the study, revised the manuscript and provided funds.

## Notes

### Competing Interest Statement

The authors have declared no competing interest.

### Author Declarations

With informed consent from the patients and approval from the hospital's ethics committee (NO. KY20212016-C-1), we recruited 66 patients at the Department of Urology, the First Affiliated Hospital of the Air Force Medical University.

