## Supplementary figures and images for "Circulating Extracellular Vesicle miRNAs for Distinguishing Prostate Cancer and Benign Prostatic Hyperplasia"

### Graphic Abstract

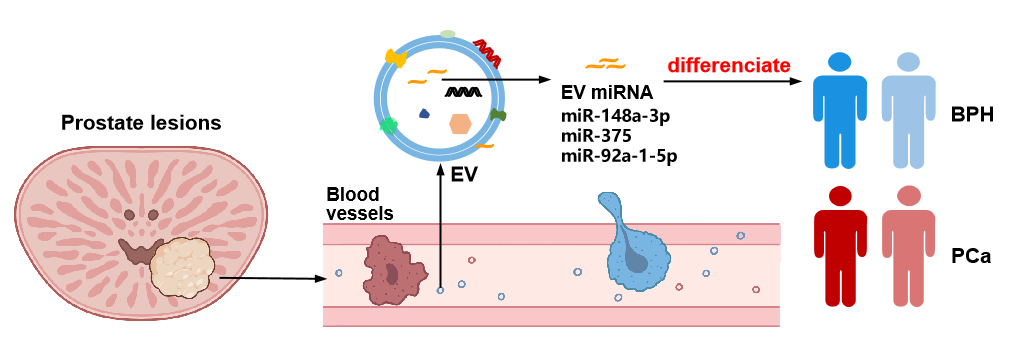

### Supplementary Figures

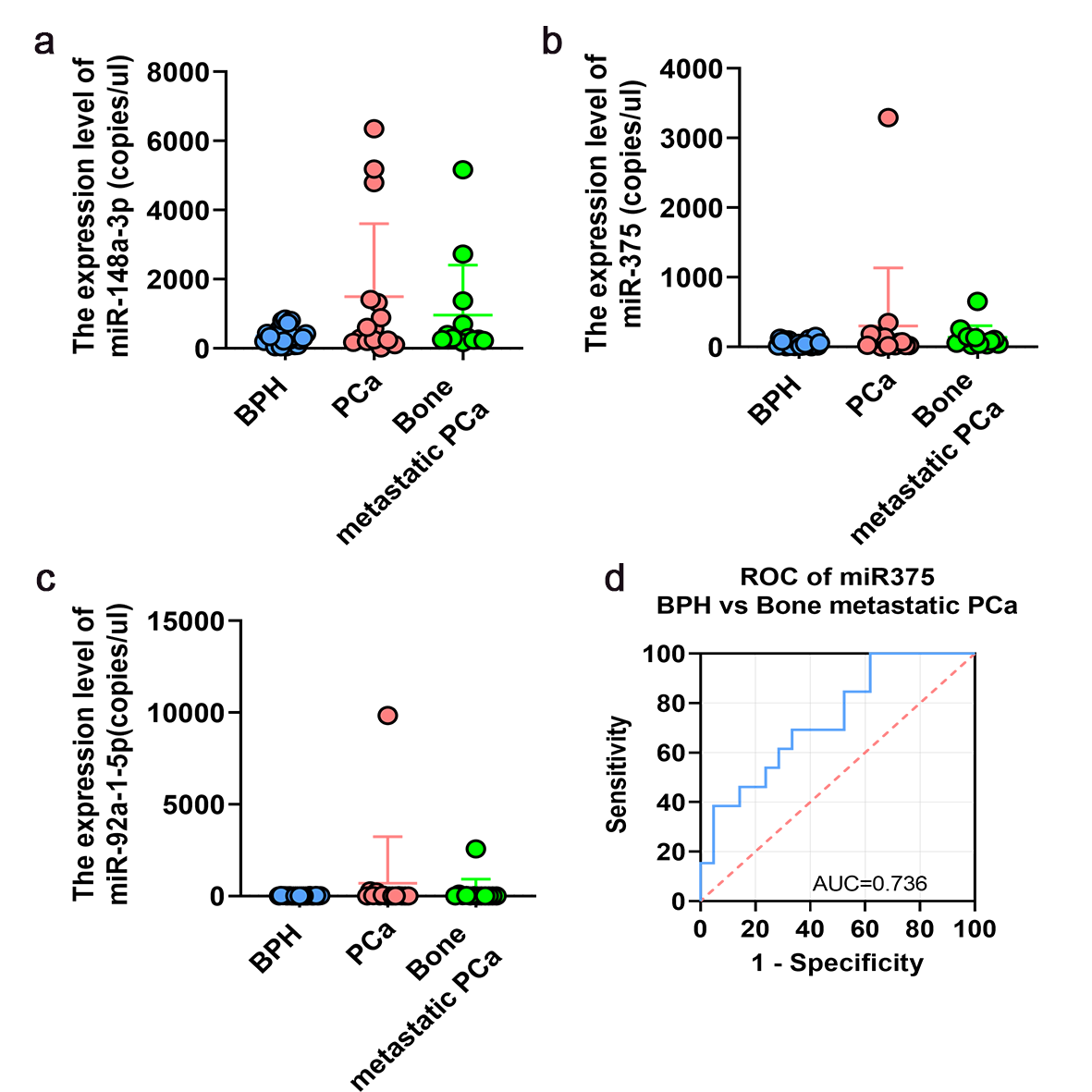
